# Development and validation of a Context-sensitive Positive Health Questionnaire (CPHQ) to measure health

**DOI:** 10.1101/2022.09.29.22280516

**Authors:** Brian M. Doornenbal, Tim van Zutphen, Rimke C. Vos, Hinke Haisma, M. Elske van den Akker-van Marle, Jessica C. Kiefte-de Jong

## Abstract

A valid context-specific measurement of health is critical for evaluating health policies and interventions. The Positive Health dialogue is a tool that is increasingly being used to evaluate health. However, this tool is meant to spark conversations about health rather than measure context-specific health. In this paper, we advance the Positive Health dialogue tool into a measurement instrument, that we denote as the Context-sensitive Positive Health Questionnaire (CPHQ). We build on previous efforts to create the CPHQ, using the Capability Approach as a theoretical framework. The measurement was developed in three stages: 1) focus groups, 2) expert consultations, and 3) validation among a representative panel of Dutch citizens. The goal of both the (1) focus groups and (2) expert consultations phases was to pilot test and refine previously proposed Positive Health questionnaires into an initial version of the CPHQ. The validation phase (3) sought to examine the initial CPHQ’s factorial validity, using Factor Analysis, and its concurrent validity, using Multivariate Regression Analysis. The developed questionnaire demonstrated adequate factorial and concurrent validity. Furthermore, it explicitly accounts for resilience, which is a key component of Positive Health. We introduced four benefits by aligning the CPHQ instrument with the Capability Approach. First, it embedded the measurement in a theoretical framework, which is required for theory development and testing. Second, it focused the questionnaire on a key concept of Positive Health - that is, on the “ability to adapt.” Third, it addressed issues of health equity by taking contextual factors into account. Fourth, it aided in developing more understandable measurement items. The introduced measurement (i.e., the CPHQ) includes 11 dimensions, which we labeled as follows: relaxation, autonomy, fitness, perceived environmental safety, exclusion, social support, financial resources, political representation, health literacy, resilience, and enjoyment.

**Highlights:**

- The Context-sensitive Positive Health Questionnaire (CPHQ) was developed using items from Positive Health questionnaires and the Capability Approach, which were then refined through focus groups and professional advice.
- The questionnaire considers individuals’ unique environment, an aspect often overlooked in health measurements that can affect how someone feels.
- By aligning the CPHQ instrument with the Capability Approach, we embedded the measurement in a theoretical framework, which is required for theory development and testing.
- By embedding the measurement in the Capability Approach and including the Brief Resilience Scale into the initial questionnaire, we accounted for a key concept of Positive Health - that is, the “ability to adapt.”
- The developed scale showed adequate factorial validity and concurrent validity.

## Introduction

Although health is regarded as an ultimate goal of medicine, measuring health is not straightforward. Good measurements measure what they purport to measure (Borsboom et al., 2004; Kelley, 1927). Clarity about the definition of health is therefore important. When it comes to measuring health, it is often unclear how health should be defined and measured (van Druten et al., 2022). How to define and measure health is more than just a theoretical issue, it has practical, policy, and healthcare implications (Jambroes et al., 2016; Leonardi, 2018). Based on the definition of health, policies and interventions are organized. Measurement instruments are needed to examine the effectiveness of these policies and interventions.

Over the years, scholars have proposed many different definitions and measurements of health (Leonardi, 2018; van Druten et al., 2022). Back in 1943, French physician Georges Canguilhem argued that health is not a fixed entity but a construct that varies depending on individual circumstances. Because of its contextual dependence, Canguilhem defined health as “the ability to adapt to one’s environment”. Five years later, in 1946, the World Health Organization (WHO) defined health as “a state of complete physical, mental and social well-being and not merely the absence of disease or infirmity”. This definition has often been discussed and criticized but has never been adapted. Particularly, the term “complete” has received criticism. The state of completeness is not measurable and out of reach for most people given the high prevalence of chronic disease (Leonardi, 2018) and the fact that health is more multidimensional and dynamic than initially assumed (Jadad & O’grady, 2008). An editorial of The Lancet (2009) reflected on the WHO definition and compared it to Canguilhem’s definition of health. It concluded that defining health as the ability to adapt - as Canguilhem proposed - will result in “a more compassionate, comforting, and creative program for medicine—one to which we can all contribute” (1943, p. 718). During a conference a few years later (Huber et al., 2011), advanced Canguilhem’s definition of health. They proposed to define health as “the ability to adapt and self-manage in the face of social, physical, and emotional challenges”.

Nowadays, the definition proposed during the conference is often denoted as Positive Health (PH; Huber et al., 2016). Governmental institutions and healthcare organizations increasingly refer to PH, among others in the Netherlands, Japan, and Iceland (Doornenbal et al., 2022). A measurement instrument for PH, which is needed for monitoring and evaluation purposes, is not yet fully developed (Doornenbal et al., 2022; Prinsen & Terwee, 2019; Van Vliet et al., 2021). For example, it is largely unclear what properties (the construct and) the measurement of PH should contain (Prinsen & Terwee, 2019). A questionnaire-based PH dialogue tool exists, but this tool was developed to inspire conversations about health during the consultation of individuals with their health and social professionals, not to measure PH. The dialogue tool is not suitable as a measurement tool given the major concerns regarding its relevance, comprehensiveness, and comprehensibility (Prinsen & Terwee, 2019). Still, more governmental institutions and healthcare organizations are accepting the definition of PH (Doornenbal et al., 2022). To help to measure health in line with the concept of PH, it is crucial to further develop and validate a suitable measurement.

To develop a measurement of PH, scholars previously tested how well the dialogue tool is suited for measuring PH. Based on tests of factorial validity, scholars turned the 42 PH items dialogue tool into a first version of a measurement model (Van Vliet et al., 2021). This measurement model (PH-17) is composed of 17 items with a six-factor structure, comprising physical fitness, mental functions, future perspective, contentment, social relations, and daily life-management. Subsequent tests explored the concurrent validity of PH-17. Whereas the 17- item questionnaire explained over 50% of the variance in measurements of overall self-rated health and happiness, it explained less than 25% of the variance in validation scales of measurements of autonomy, personal growth, stability, and self-care (Doornenbal et al., 2022). Philippens and colleagues (2021) provided support for construct validity by finding a positive impact of a combined lifestyle intervention on PH-17. Concerns remain about the fit between the PH measurement and what the PH measurement purports to measure (Doornenbal et al., 2022; Prinsen & Terwee, 2019). These concerns are caused by among others clarity about whether the construct of PH refers to “patients’ experiences or perceptions with different aspects of health, their level of (aspects of) health and/or their satisfaction with (aspects of) health” (Prinsen & Terwee, 2019, p. 75). Further, the measurement instrument should be more inclusive toward vulnerable populations for which contextual factors (e.g., neighborhood adversity, perceptions of discrimination, and social resources) can hamper coping and recovery processes (de Groot et al., 2019).

In this paper, we further develop and test a context-sensitive measurement of PH, which we denote in this paper as Context-specific Positive Health (CPH). We partly take an empirical approach to test the validity of the measurement, similar to previous efforts (Doornenbal et al., 2022; Van Vliet et al., 2021). Beyond this empirical approach, we take a theoretical approach to address concerns about construct clarity (Prinsen & Terwee, 2019; Van Vliet et al., 2021) and account for contextual dependence of health. We align the measurement of CPH to the Capability Approach (Chiappero-Martinetti & Venkatapuram, 2014; Nussbaum, 2008; Sen, 1990), which has four major benefits. First, it helps to embed the construct of CPH in a theoretical framework - which is needed for theory building and testing (Prinsen & Terwee, 2019). Second, it creates clarity about the focus of CPH - that is, on the “ability to adapt”. Third, it addresses issues of health equity (de Groot et al., 2019). Fourth, it provides a structure for creating more comprehensible measurement items - which scholars called for (Prinsen & Terwee, 2019).

## Methods

The Context-sensitive Positive Health Questionnaire (CPHQ) was developed in three phases: 1) focus groups, 2) expert consultations, and 3) validation in a representative panel of citizens. As input for these phases, a questionnaire was used. Both the (1) focus groups and (2) expert consultations phase were intended to pilot test and refine this questionnaire with both public health experts (e.g., specialized in poverty) and citizens from different backgrounds (e.g., educational, cultural, health conditions). The goal of the validation phase (3) was to examine the factorial validity and concurrent validity of the refined questionnaire.

The questionnaire that served as a starting point for the three phases was inspired by the Positive Health dialogue tool (Huber et al., 2016). The Positive Health dialogue tool consists of 42 statements, each belonging to one of six dimensions, initially named: bodily functions, mental functions and perception, spiritual existential dimension, quality of life, social and societal participation, and daily functioning (Van Vliet et al., 2021). The 42 PH statements were extended with items related to Nussbaum’s 10 core capabilities for adult-wellbeing (Nussbaum, 2011), focusing on the context of individuals. We developed the context items on existing validated quality of life questionnaires and other context-sensitive sources such as the resilience monitor, self-sufficiency matrix, monitor broad welfare, and livability index (Mink et al., 2015). These items were divided into interpersonal context (16 items), social context (24 items), and environmental context (14 items). Specific contexts can function as conversion factors that either support or hinder people’s capabilities (Chiappero-Martinetti & Venkatapuram, 2014). Examples of conversion factors are gender, ethnicity, culture, and laws and regulations (Chiappero-Martinetti & Venkatapuram, 2014). The initial questionnaire items were formulated in line with the Capability Approach (Nussbaum, 2011; Sen, 1990). We rephrased items such that they focused on endowments, capabilities, or functionings rather than states. For example, “I know what I can and what I can’t” was rephrased as “I am able to perform tasks and activities adequately.” Based on the focus groups and expert input, the questionnaire was extended with items that were considered to be missing by the citizens and/or experts which were related to resilience, social support, relaxation, and autonomy.

For this study, ethical approval was obtained from the Medical Ethical Review Board of the Leiden University Medical Center (protocol 19-035) and the Research Ethics Committee of the Faculty of Spatial Sciences of the University of Groningen (protocol 202007).

### 1. Focus groups

The focus groups were aimed to assess the relevance and comprehensibility of the 42 PH-items as well as the contextual items about perceived health. As a preparation for the focus group participants were asked to fill in the 42 PH-items. During the focus groups, the participants were first asked how they defined perceived health in their own words. Next, all domains of the 42 PH-items conversation tool were discussed if they were relevant in light of their definition of perceived health, the comprehensibility, and whether items were missing. Because resilience was previously mentioned as a core element in the original definition of Huber who proposes to see health more as a power than a state, defined as the power to be resilient (2011) and previous research showed that the initial PH scales explained little variation in resilience (Doornenbal et al., 2022), we specifically asked citizens whether resilience was sufficiently reflected and whether it would be of added value to add one or more other ‘potential’ items to the PH model. For the contextual items, the output from the focus groups was coded and analyzed according to the endowments, capabilities, and conversion factors from the Capability Approach. We included participants distributed over various age groups, gender, health conditions (with or without chronic disease), socioeconomic background, and cultural backgrounds. In total, we organized six focus group sessions, each with five participants, which appeared to be sufficient to achieve saturation.

### 2. Expert consultations

Next to the focus groups, experts from the medical, policy, and research domains were consulted on the definition of health and the domains and items belonging to this definition. Through a questionnaire, the experts were asked to mention stronger and weaker points of the definition of PH. After that, they could indicate and rank the most important domains of health from a list with domains from different questionnaires, such as PH-42 (Van Vliet et al., 2021), EQ-5D (Herdman et al., 2011), ICECAP-A (Al-Janabi et al., 2012), and HR-SWB (de Vries et al., 2016). Also, new domains could be listed. Subsequently, the individual items of the PH were shown for which respondents could indicate which of them belong to health. The goal of the expert consultation was to further refine the questionnaire before empirically testing the factorial validity and concurrent validity.

Furthermore, experts with first-hand poverty experience indicated that they were hesitant to complete the questionnaire because some items did not apply to their situation. Based on this feedback, the language was adjusted. For example, positively phrased questions such as “I do not have financial problems” were rephrased as “I have debts” to better reflect a low socioeconomic situation. Furthermore, items about the personal, social and environmental context were confirmed to be relevant to add to the 42-item PH questionnaire. In total, the 42 items were complemented with 12 items on autonomy, relaxation, resilience, and social support to a total of 54 items developed and formulated based on the Capability Approach (See Supplemental Table 1).

### 3. Validation in a representative panel of citizens

The goal of the validation phase was to examine the factorial validity and concurrent validity of the refined questionnaire. An Exploratory Factor Analysis (EFA), followed by a Confirmatory Factor Analysis (CFA) was conducted to test the factorial validity—that is, the extent to which a putative structure of a scale is recoverable in a set of test scores. Data used for these analyses were gathered through a Dutch independent Internet panel Flycather, which operates in line with ISO standards. In total, 1002 panel members, representative of the Dutch population in terms of sociodemographic background, participated in this study. The data collection took place in December 2020.

The measurement model extracted during the Factor Analysis (FA) - the adapted CPHQ was compared with other health scales to assess the concurrent validity—that is, the degree to which a new test compares to an established test. For this, participants were asked to fill out the adapted CPHQ (54 items) and the following validation scales:

#### Brief resilience scale (BRS)

To measure resilience, three positive-worded items of the brief resilience scale (BRS) were used (Smith et al., 2008). The items used were: (1) “I tend to bounce back quickly after hard times”, (2) “It does not take me long to recover from a stressful event”, and (3) “I usually come through difficult times with little trouble”. Answers were given on a 5-point Likert scale ranging from *strongly disagree* (=1) to *strongly agree* (=5).

#### Experienced Lifestyle Behaviors (SLB-4D)

Lifestyle behaviors were measured on four dimensions: physical activity, healthy eating, substance use, and rest. These four dimensions are systematically found to be related to health (Downes et al., 2021; Noble et al., 2015). To assess the dimensions, direct measurements were used: “… is going the way I want it to”. Participants were asked to respond on a 5-point Likert scale ranging from *strongly disagree* (=1) to *strongly agree* (=5). We refer to this measurement as the SLB-4D (Salut Lifestyle Behaviors four-Dimensions).

#### Health-related subjective well-being (HR-SWB)

Health-related subjective well-being (HR-SWB) was measured using the measurement proposed by De Vries and colleagues (2016). This measurement comprises five dimensions: (1) bodily independence, (2) happiness, (3) loneliness, (4) autonomy, and (5) personal growth. Each domain was measured using one item, such as “I feel lonely” (loneliness). Responses were rated on a 5-point Likert scale ranging from *strongly disagree* (= 1) to *strongly agree* (= 5).

#### EuroQol five-Dimensions (EQ-5D)

The EQ-5D-5L (EuroQol five-Dimensions) captures the following domains of health: mobility, self-care, usual activities, pain/discomfort, and anxiety/depression. Answer categories of questions on these were assessed on a 5-point Likert scale. At last, a visual analog scale was used to measure the overall self-rated health of the respondent that day (Herdman et al., 2011).

#### ICEpop CAPability measure for Adults (ICECAP-A)

The ICECAP-A (ICEpop CAPability measure for Adults) was used to measure well-being following the Capability approach in terms of individuals’ capabilities (Al-Janabi et al., 2012). The measurement comprises five domains: stability, attachment, autonomy, achievement, and enjoyment. Each of these domains was measured using one statement on a 4-point scale.

### Analyses

For the analyses, the data (n = 1002) were randomly and evenly partitioned into two datasets: a training and a test dataset. The training dataset was used for the Exploratory Factor analysis (EFA), whereas the test dataset was used for the CFA. Splitting the data into a training and test dataset helps to evaluate how well unknown data fit the measurement model.

To examine the dimensionality of the data, a series of factor models were fitted. We began with a one-factor model and incrementally added one factor (k + 1) at a time. While fitting the models, all items were allowed to load on all the factors on the model – no a priori restrictions were imposed on the factorial structure. Factors were added to the model until the model (1) demonstrated adequate goodness of fit in terms of the Comparative Fit Index (CFI) and Tucker-Lewis Index (TLI), the Root Mean Square Error of Approximation (RMSEA), and the Standardized Root Mean Square Residual (SRMR); (2) explained most of the variance; (3) had an interpretable structure in which at least two items load strongly (i.e., ≥ 0.40) on each factor only (i.e., no cross-loadings allowed).

The goodness of fit was assessed using CFA with robust Maximum Likelihood (MLR). Compared to Maximum Likelihood (ML) estimation, MLR is less dependent on the assumption of multivariate normal distribution (Li, 2016). To compute the goodness of fit indices (i.e., CFI, TLI, RMSEA, and SRMSR), items were selected during the EFA. The three items with the highest factor loadings (≥ 0.40) and without cross-loadings were selected to compute the goodness of fit indices.

While exploring the factor structure, Horn’s parallel analysis was applied (Hayton et al., 2004; Horn, 1965) to limit our search to dimensionalities for which the likelihood is greater than random chance. Specifically, the *k*th eigenvalue of the sample covariance was compared with the sampling distribution of the *k*th eigenvalue obtained through Monte Carlo simulation from random independent data. Only the factor structures for which the *k*th eigenvalue of the sample data is substantially larger than the *k*th eigenvalue of the simulated data have a dimensionality that is greater than one would expect by random chance.

To evaluate the relationship between CPHQ and other measurements of health (i.e., BRS, SLB-4D, HR-SWB, EQ-5D, ICECAP-A), multivariate regression analyses were conducted. During each regression analysis, the factors of the final measurement model of CPHQ were used as independent variables, whereas the other measurements of health (incl. underlying domains) were each time used as a dependent variable. The proportion of the variance for the dependent variable that is explained by the independent variables in the regression models (R^2^) was used as a statistical measure that represents the strength of the statistical relationships between CPHQ and the other health domains—which we denote as validation scales. This analysis was intended to assess the concurrent validity.

## Results

Inspection of data suggested that the training dataset was suitable for EFA. The adequacy of the sample size for the EFA (n = 501) was “very good” (Comfrey & Lee, 1992, p. 217), with a subject-to-item ratio of 4.5:1. The KMO test yielded a statistic of 0.95, suggesting the data set contains a significant proportion of variance among variables that might be common variance (caused by underlying factors). Bartlett’s test of sphericity yielded significant results, χ2(111)=3095.04, p<0.001, implying that the data are suitable for performing factor analysis because the correlations among variables are greater than one would expect by chance.

Using the training data to explore the factor structure, we limited the search to 15 factors because Horn’s parallel analysis (Hayton et al., 2004; Horn, 1965) suggested that a dimensionality of more than 15 factors is unlikely compared to the dimensionality expected by random chance. This search was further narrowed down to 11 dimensions, because for 12 dimensions and more, at least one dimension did not have strong item loadings (i.e., exceeding 0.40). As reported in Supplemental Table 2, most variance (> 50%) was explained when the data were structured into 9 dimensions or more. Thus, we focused on factor structures ranging from 9 to 11 dimensions.

During the subsequent CFA, the items showed positive factor loadings on the respective domains with an average standardized coefficient of 0.793, ranging from 0.543 to 0.900 (Table 2). Thus, an 11 dimensions factor model adequately described the data. Across the factor structures ranging from 9 to 11 dimensions, a 11-dimensions factor structure had the best goodness of fit indices (CFI = 0.944, TLI = 0.932, RMSEA = 0.049, SRMR = 0.050). The goodness of fit indices were computed based on the test data (n = 501) – i.e., the data that were not used during the EFA. The 11 dimensions solution had an interpretable factor structure in which at least two items load strongly (i.e., ≥ 0.40) on each factor only during the EFA (Table 1).

**Table 1.**
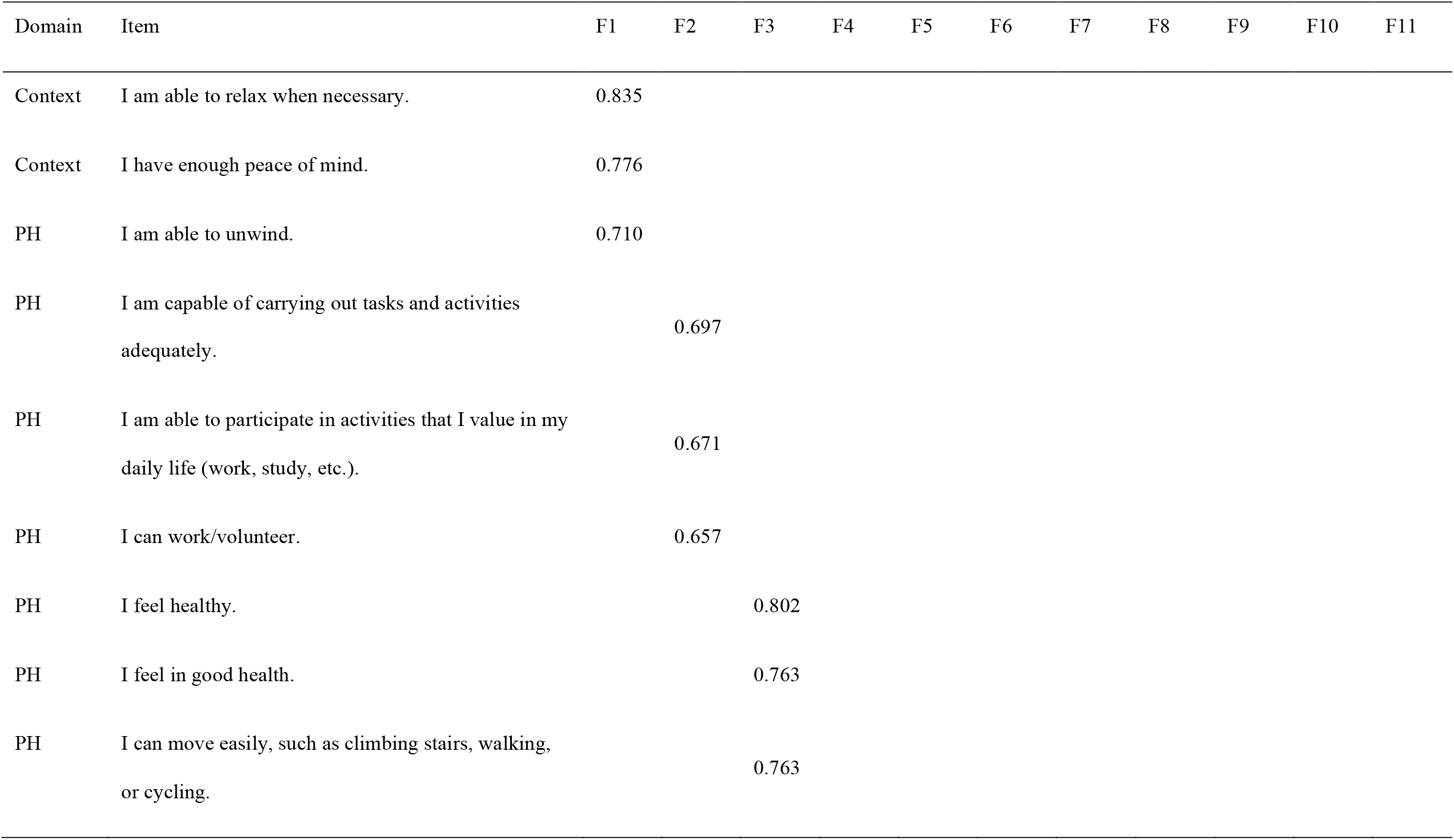
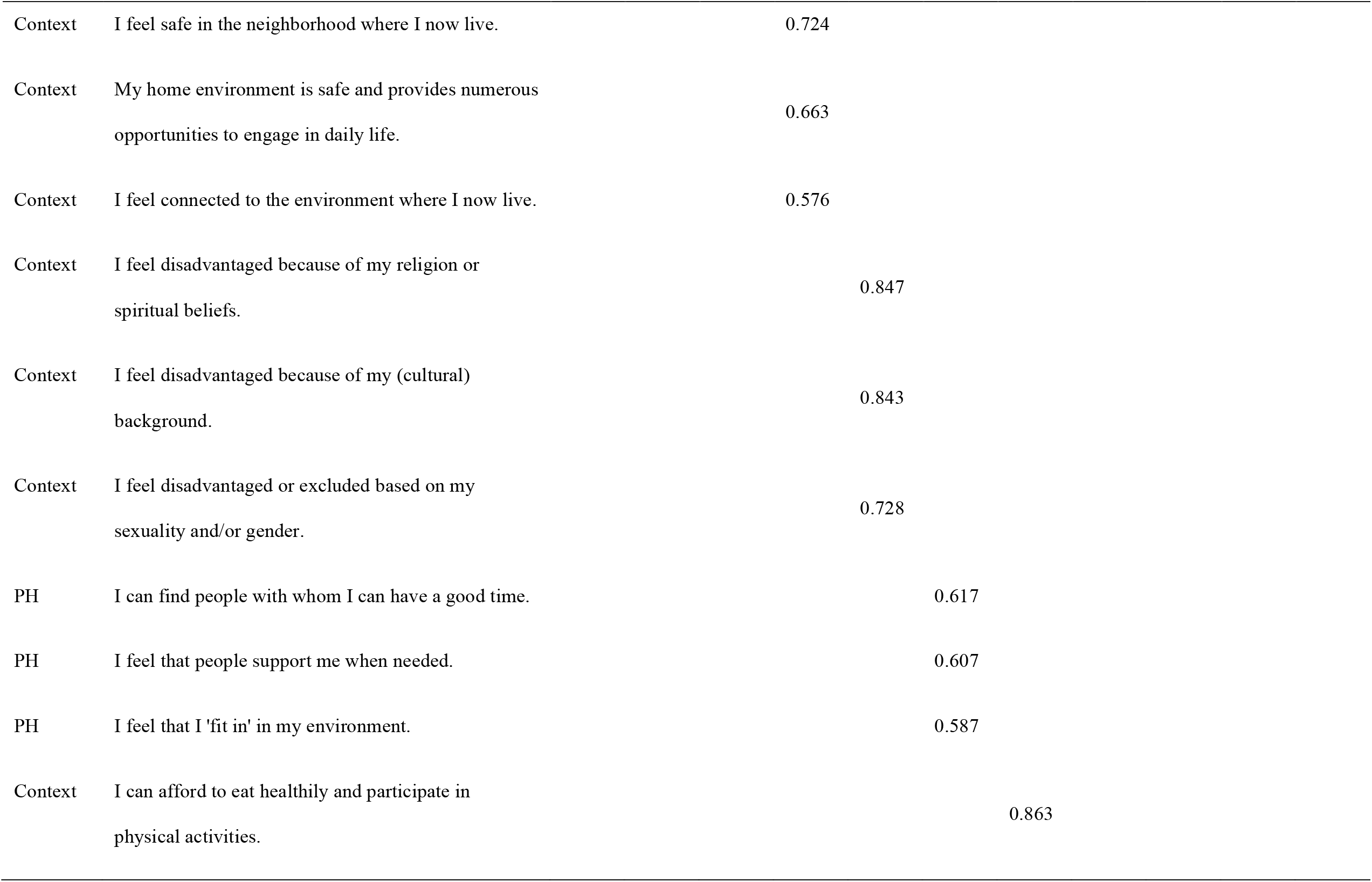
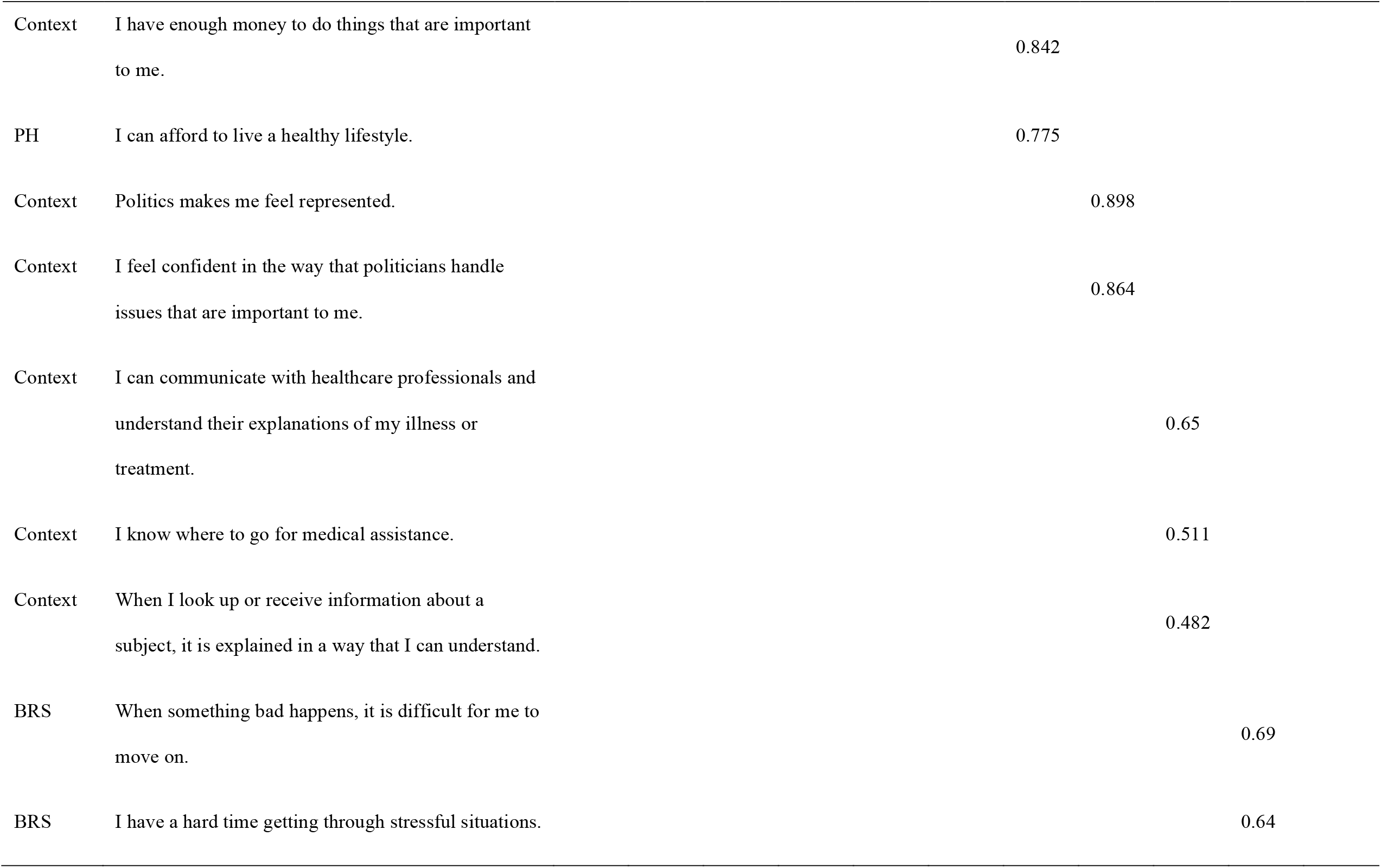
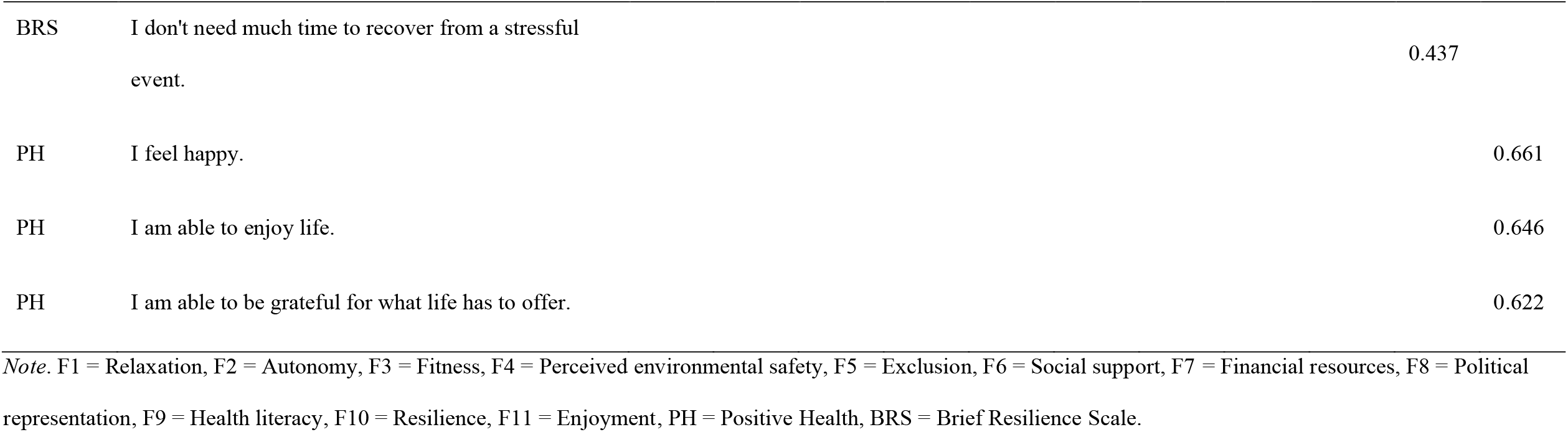
Standardized weights of the exploratory eleven-factor model using oblimin rotated factors.

**Table 2.** Parameter estimates Confirmatory Factor Analysis (CFA) using robust Maximum Likelihood (MLR)

| Latent<br>Factor | Description | **B** | **SE** | **Z** | **$\beta$** | ***p*** |
| --- | --- | --- | --- | --- | --- | --- |
| F1 | I am able to relax when necessary. | 1.000 |  |  | 0.894 |  |
| F1 | I have enough peace of mind. | 1.046 | 0.045 | 23.254 | 0.817 | *** |
| F1 | I am able to unwind. | 0.785 | 0.042 | 18.670 | 0.788 | *** |
| F2 | I am capable of carrying out tasks and activities adequately. | 1.000 |  |  | 0.785 |  |
| F2 | I am able to participate in activities that I value in my daily life (work, study, etc.). | 1.351 | 0.096 | 14.090 | 0.881 | *** |
| F2 | I can work/volunteer. | 1.292 | 0.116 | 11.128 | 0.689 | *** |
| F3 | I feel healthy. | 1.000 |  |  | 0.890 |  |
| F3 | I feel in good health. | 1.075 | 0.043 | 25.022 | 0.873 | *** |
| F3 | I can move easily, such as climbing stairs, walking, or cycling. | 0.899 | 0.064 | 14.152 | 0.660 | *** |
| F4 | I feel safe in the neighborhood where I now live. | 1.000 |  |  | 0.845 |  |
| F4 | My home environment is safe and provides numerous opportunities to engage in daily life. | 0.725 | 0.080 | 9.089 | 0.644 | *** |
| F4 | I feel connected to the environment where I now live. | 0.916 | 0.102 | 8.976 | 0.582 | *** |
| F5 | I feel disadvantaged because of my religion or spiritual beliefs. | 1.000 |  |  | 0.870 |  |
| F5 | I feel disadvantaged because of my (cultural) background. | 1.009 | 0.051 | 19.755 | 0.890 | *** |
| F5 | I feel disadvantaged or excluded based on my sexuality and/or gender. | 0.900 | 0.061 | 14.795 | 0.738 | *** |
| F6 | I can find people with whom I can have a good time. | 1.000 |  |  | 0.753 |  |
| F6 | I feel that people support me when needed. | 0.965 | 0.078 | 12.368 | 0.803 | *** |
| F6 | I feel that I 'fit in' in my environment. | 1.101 | 0.073 | 14.995 | 0.814 | *** |
| F7 | I can afford to eat healthily and participate in physical activities. | 1.000 |  |  | 0.881 |  |
| F7 | I have enough money to do things that are important to me. | 1.155 | 0.052 | 22.115 | 0.854 | *** |
| F7 | I can afford to live a healthy lifestyle. | 0.889 | 0.057 | 15.580 | 0.805 | *** |
| F8 | Politics makes me feel represented. | 1.000 |  |  | 0.888 |  |
| F8 | I feel confident in the way that politicians handle issues that are important to me. | 1.014 | 0.099 | 10.199 | 0.900 | *** |
| F9 | I can communicate with healthcare professionals and understand their explanations of my illness or treatment. | 1.000 |  |  | 0.812 |  |
| F9 | I know where to go for medical assistance. | 0.887 | 0.078 | 11.342 | 0.776 | *** |
| F9 | When I look up or receive information about a subject, it is explained in a way that I can understand. | 0.682 | 0.081 | 8.444 | 0.543 | *** |
| F10 | When something bad happens, it is difficult for me to move on. | 1.000 |  |  | 0.829 |  |
| F10 | I have a hard time getting through stressful situations. | 1.008 | 0.054 | 18.804 | 0.782 | *** |
| F10 | I don't need much time to recover from a stressful event. | 0.713 | 0.078 | 9.179 | 0.614 | *** |
| F11 | I feel happy. | 1.000 |  |  | 0.880 |  |
| F11 | I am able to enjoy life. | 0.918 | 0.039 | 23.694 | 0.888 | *** |
| F11 | I am able to be grateful for what life has to offer. | 0.679 | 0.051 | 13.225 | 0.696 | *** |

*Note.* \*\*\* = $p < .001$ ; B = unstandardized estimates; SE = standardized error; F1 = Relaxation, F2 = Autonomy, F3 = Fitness, F4 = Perceived environmental safety, F5 =
Exclusion, F6 = Social support, F7 = Financial resources, F8 = Political representation, F9 = Health literacy, F10 = Resilience, F11 = Enjoyment.

The factors of the 11 dimensions structure are sufficiently distinct. The correlations between the factors ranged from .074 (Exclusion and Political representation) to .631 (Relaxation and Enjoyment). In addition to the support for the discriminant validity, the interrelatedness amongst individual items within a factor was sufficient. The Cronbach alpha’s of the factors ranged from .71 (Perceived environmental safety) to .89 (Political representation).

To examine the concurrent validity, the relationships between the 11-factor model and each of the validation scales were tested (Table 3). Testing the multiple correlation coefficient (i.e., R^2^) between the 11-factor model and the validation scales, we found relationships of mixed strength. The 11-factor model explained less than 25% of the variance of some of the validation scales: *substance use* (SLB-4D, R^2^ = 0.08), *self-care* (EQ-5D, R^2^ = 0.12), *personal growth* (HR-SWB, *R*^2^ = 0.12), *autonomy* (HR-SWB, *R*^2^ = 0.19; ICECAP-A, *R*^2^ = 0.17), and *stability* (ICECAP-A, *R*^2^ = 0.23). Between 25% and 40% was explained by the model in validation scales: *physical independence* (HR-SWB, *R*^*2*^ = 0.26), *healthy eating* (SLB-4D, *R*^2^ = 0.27), *attachment* (ICECAP-A, *R*^2^ = 0.29), *achievement* (ICECAP-A, *R*^2^ = 0.33), *physical activity* (SLB-4D, *R*^2^ = 0.35), *pain/discomfort* (EQ-5D, *R*^2^ = 0.36), and *mobility* (EQ-5D, *R*^2^ = 0.39). The model explained more than 40% of the variance in validation scales: *usual activities* (EQ-5D, *R*^2^ = 0.42), *loneliness* (HR-SWB, *R*^2^ = 0.43), *enjoyment* (ICECAP-A, *R*^2^ = 0.44), *EQ-VAS* (EQ-5D, *R*^2^ = 0.45), *anxiety/depression* (EQ-5D, *R*^2^ = 0.49), *rest* (SLB-4D, *R*^2^ = 0.54), *resilience* (BRS, *R*^2^ = 0.57), and *happiness* (HR-SWB, *R*^2^ = 0.83).

**Table 3.** Multivariate regression analyses between the 11 factors and validation scales.

| Scale | Domain | Standardized loadings ($\beta$) on Positive Health dimensions | | | | | | | | | | | $R^2$ |
| --- | --- | --- | --- | --- | --- | --- | --- | --- | --- | --- | --- | --- | --- |
|  |  | F1 | F2 | F3 | F4 | F5 | F6 | F7 | F8 | F9 | F10 | F11 |  |
| BRS | Resilience | 0.195**<br>* | 0.062 | 0.018 | -0.002 | 0.011 | 0.065 | 0.008 | 0.065* | 0.034 | 0.755*<br>** | 0.077 | 0.567 |
| SLB-4D | Physical activity | 0.157** | 0.052 | 0.348**<br>* | 0.037 | -0.016 | -0.021 | 0.128*<br>* | 0.005 | -0.011 | -0.023 | 0.088 | 0.350 |
|  | Healthy eating | 0.268**<br>* | -0.039 | 0.149** | 0.052 | 0.004 | 0.122* | 0.175*<br>** | -0.104* | 0.077 | -0.002 | -0.015 | 0.267 |
|  | Substance use | 0.042 | 0.042 | 0.075 | 0.076 | 0.060 | -0.093 | 0.012 | -0.022 | 0.142** | -0.030 | 0.094 | 0.078 |
|  | Rest | 0.651**<br>* | -0.025 | 0.025 | 0.108** | 0.037 | 0.036 | -0.001 | 0.022 | -0.018 | 0.002 | 0.026 | 0.544 |
| HR-SWB | Physical independence | 0.006 | 0.173** | 0.402**<br>* | 0.062 | 0.028 | -<br>0.102* | 0.028 | -0.059 | -0.005 | -0.029 | 0.007 | 0.262 |
|  | Happiness | 0.081** | -0.019 | -0.015 | -0.003 | -0.022 | 0.063*<br>* | 0.020 | -0.005 | -<br>0.064** | -0.006 | 0.858**<br>* | 0.834 |
|  | Loneliness | 0.297**<br>* | -0.025 | -0.033 | 0.021 | 0.114*<br>** | 0.251*<br>** | 0.073 | -0.054 | -0.012 | -0.065 | 0.177** | 0.427 |
|  | Autonomy | 0.206**<br>* | 0.166** | -0.122* | 0.037 | 0.015 | 0.108* | 0.044 | -0.043 | 0.158** | 0.010 | 0.024 | 0.194 |
|  | Personal growth | -<br>0.207**<br>* | 0.078 | 0.073 | -0.011 | -0.003 | 0.001 | -0.128* | 0.082 | 0.205**<br>* | 0.023 | 0.249**<br>* | 0.118 |
| EQ-5D | Mobility | -0.001 | 0.232**<br>* | 0.563**<br>* | -0.092* | 0.08* | -0.058 | -0.038 | 0.056 | -0.075 | -0.013 | -0.129* | 0.392 |
|  | Self-care | -0.080 | 0.203** | 0.183** | -0.017 | 0.085 | -0.076 | -0.020 | 0.010 | 0.005 | 0.034 | 0.106 | 0.115 |
|  | Usual activities | 0.017 | 0.385**<br>* | 0.379**<br>* | -0.066 | -<br>0.077* | -0.069 | 0.108* | 0.025 | -0.058 | -0.037 | -0.074 | 0.416 |
|  | Pain/Discomfort | 0.106* | 0.130* | 0.525**<br>* | -0.100* | -0.017 | -0.045 | 0.085 | 0.061 | -0.055 | -0.025 | -0.125* | 0.361 |
|  | Anxiety/Depression | 0.397**<br>* | 0.013 | -0.009 | -0.070 | -<br>0.129*<br>** | 0.186*<br>** | 0.064 | 0.005 | -0.082* | -<br>0.175*<br>** | 0.216**<br>* | 0.489 |
|  | EQ-VAS | 0.152** | 0.122* | 0.409**<br>* | -0.030 | -0.010 | -0.008 | 0.071 | 0.006 | -0.020 | -0.033 | 0.118* | 0.446 |
| ICECAP-A | Stability | 0.278**<br>* | 0.131* | -0.036 | 0.081 | -0.081 | 0.085 | 0.008 | 0.035 | 0.002 | -0.072 | 0.061 | 0.234 |
|  | Attachment | 0.065 | 0.021 | -0.047 | 0.049 | 0.038 | 0.250*<br>** | -0.065 | 0.055 | -0.058 | -0.002 | 0.328**<br>* | 0.293 |
|  | Autonomy | 0.125* | 0.296**<br>* | 0.083 | 0.015 | -0.034 | 0.061 | 0.021 | -0.016 | 0.029 | -0.075 | -0.028 | 0.208 |
|  | Achievement | 0.097 | 0.347**<br>* | 0.111* | -0.009 | -<br>0.116*<br>* | 0.074 | 0.016 | 0.005 | -0.011 | 0.020 | 0.123* | 0.332 |
|  | Enjoyment | 0.224**<br>* | 0.169** | -0.005 | 0.032 | -<br>0.097*<br>* | 0.180*<br>** | -0.044 | 0.037 | -0.056 | 0.026 | 0.300**<br>* | 0.441 |

*Note.* $N = 501$ ; $R^2$ denotes the amount of variance explained by the six domains of Positive Health; F1 = Relaxation, F2 = Autonomy, F3 = Fitness, F4 = Perceived environmental safety, F5 = Exclusion, F6 = Social support, F7 = Financial resources, F8 = Political representation, F9 = Health literacy, F10 = Resilience, F11 = Enjoyment; *BRS* = Brief Resilience Scale; *SLB-4D* = Salut Lifestyle Behavior 4-Dimensional; *HR-SWB* = Health-Related Subjective Well-Being; *EQ-5D* = EuroQol 5-Dimensional; *EQ-VAS* = EuroQol Visual Analog Scale; *ICECAP-A* = ICEpop CAPability measure for Adults.
\* = $p < .05$ .
\*\* = $p < .01$ .
\*\*\* = $p < .001$ .

Testing the relationship between the 11 factors and the validation scales, we found that *all* factors were statistically important in explaining variance across the validation scales (Table 3). However, five factors (i.e., F1, Autonomy, Fitness, Resilience, Enjoyment) were well covered by the validation scales. For example, *F1* had a strong statistical significant association with *rest* (SLB-4D, β = 0.651, *p* < 0.001) and *anxiety/depression* (EQ-5D, β = 0.397, *p* < 0.001); *Autonomy* was an important predictor of *usual activities* (EQ-5D, β = 0.385, *p* < 0.001) and *achievement* (ICECAP-A, β = 0.347, *p* < 0.001); *Fitness* was strongly related to among others *mobility* (EQ-5D, β = 0. 563, *p* < 0.001), *pain/discomfort* (EQ-5D, β = 0.563, *p* < 0.001), *physical independence* (HR-SWB, β = 0. 402, *p* < 0.001), and *physical activity* (SLB-4D, β = 0.348, *p* < 0.001); *Resilience* had also a strong statistical relationship with the *Brief Resilience Scale* (BRS, β = 0.755, *p* < 0.001); and *Enjoyment* was an important predictor of *happiness* (HR-SWB, β = 0.858, *p* < 0.001). The remaining six factors (i.e., Perceived environmental safety to Health literacy) were statistically important in explaining variance in the validation scales but less clearly related to one of the validation scales.

## Discussion

This study aimed to develop a Context-sensitive Positive Health Questionnaire (CPHQ), extending the previous efforts to develop a Positive Health (PH) measurement model. Compared to the PH measurement model, the CPHQ includes context items following the constitutive elements of the Capability Approach (Chiappero-Martinetti & Venkatapuram, 2014; see Supplemental Table 1). The Capability Approach served as a theoretical framework, responding to calls for: a theoretical framework needed to build and test theory (Doornenbal et al., 2022; Prinsen & Terwee, 2019), clarity about the focus of CPHQ (on the “ability to adapt”), accounting for health inequality (de Groot et al., 2019), and more comprehensible measurement items (Prinsen & Terwee, 2019). While developing CPHQ, input from citizens and professionals on positive health and context items was included to account for content validity. Factor Analysis and Regression analysis were conducted to assess the factorial validity and concurrent validity.

An initial questionnaire, which was developed based on PH and the Capability Approach, was refined during focus discussions and expert consultation. The refined questionnaire contained PH items and items related to resilience, social support, relaxation, and autonomy. This refined questionnaire was used during a Factor Analysis, for which data were gathered among a representative panel of Dutch citizens. The Exploratory Factor Analysis (EFA) suggested a model containing 11 dimensions, which we labeled as: relaxation, autonomy, fitness, perceived environmental safety, exclusion, social support, financial resources, political representation, health literacy, resilience, and enjoyment. The found factors partly overlap with the initial PH questionnaire, which contains the dimensions: physical fitness, mental functions, future perspective, contentment, social relations, and daily life-management (Van Vliet et al., 2021). Hence, the CPHQ seems advances the PH, responding to the call from our participants of the focus groups and expert consultation. The integration of the Capability Approach responds to the call for a theoretical framework for PH (Doornenbal et al., 2022; Prinsen & Terwee, 2019). Support for the factorial validity of the 11-dimensional CPHQ was found through Confirmatory Factor Analysis (CFA).

The tests of concurrent validity showed that all 11 dimensions of the CPHQ were statistically important in explaining variance across the validation scales. Five factors were well covered by the validation scales. In particular, these factors showed a strong relationship with *Anxiety/Depression, Achievement, Mobility, Pain/Discomfort, Physical independence, Resilience*, and *Happiness*. The remaining factors had significant but weak relationships with the validation scales. The factors that showed weaker relationships with the validation scales were mostly the newly added items on context (i.e., perceived environmental safety, social support, exclusion, financial resources, health literacy, and political representation), which were formulated in line with the principles of the Capability Approach (Chiappero-Martinetti & Venkatapuram, 2014). Possibly, these factors affect the extent to which persons can feel well, as we will discuss next.

Applying the Capability Approach as a framework for the CPHQ helps to focus on how health, as a functioning, can be achieved by analyzing a person’s resources as well as (conversion factors) that could influence the transformation of such resources into health capabilities. For example, *Perceived environmental safety* was characterized by contextual items related to connectedness and feeling safe in one’s environment and was significantly related to the *Mobility* and the *Pain/Discomfort* domain of the EQ-5D as well as to *Rest* (SLB-4D). A safe environment is a resource that impacts the ability to control one’s life (capability) as well as impact actual behavior such as physical activity (functionings) which may explain the specific link between mobility and pain (Won et al., 2016). However, as we did not find a strong correlation with other validation scales, this factor may represent a distinct environmental stressor related to perception instead of actual exposure and act more as a personal conversion factor that impacts mobility decisions (e.g., a chosen mode of transportation), for example, due to noise and traffic pollution and dissatisfaction related to relaxation (Marquart et al., 2021). *Exclusion* on the other hand was significantly but weakly related to *Anxiety/Depression, Achievements*, and *Loneliness* which was in the expected direction as discrimination have been found to affect mental health (Murney et al., 2020) as well as human capital (Caputo, 2002). In this situation, the domain *Exclusion* can act as a conversion factor whereas the domain *Social support*, which was related to the items *Loneliness* (HR-SWB), *Attachment* (ICECAP-A), and *Anxiety/Depression* (EQ-5D), included items such as “feeling supported if needed” and “ability to find people to engage with” which combines personal conversion factor and capabilities.

Similarly, the factor *Financial resources* was characterized by having sufficient financial resources to live a healthy life and had a weak statistical significant relationship with *Personal growth* (HR-SWB) and *Usual activities* (EQ-5D) as well as *Physical activity* and *Healthy eating* (SLB-4D). Financial hardship occurs when one has insufficient financial resources to adequately meet a household’s needs. Experiencing this type of deprivation can impact health and well-being by inducing psychological distress, lack of access to health-promoting resources such as sports and healthy food as well as little participation in leisure activities (Tucker-Seeley et al., 2013).

At last, both *Health literacy* and *Political representation* did not show a strong relationship with the validation scales, which may be explained by the fact that none of the validation scales adequately captured these domains. However, in light of the Capability Approach, both *Health literacy* and *Political representation* are relevant to consider when measuring health. For example, previous work by Pithara (2020) has re-conceptualized health literacy by using the Capability Approach in which the authors highlight the need for addressing health literacy as a capability alongside other health-promoting factors instead of focusing on narrow competency-related goals that are mostly used in health literacy measurement scales (Nguyen et al., 2015). Indeed, the current CPHQ represents a combination of health literacy factors combining the ability to communicate with health professionals as well as knowing where to go for medical support and understanding medical information. Also, the importance of political representation has been acknowledged in the Capability Approach of human well-being as political representation is an important conversion factor for access to health-promoting resources (Robeyns, 2005).

Overall, the CPHQ measurement model showed a higher explained variance in resilience and mental health indicators (i.e., Happiness and Anxiety/Depression) and was similar in explaining the other validation scales when compared to the previous PH scale (Doornenbal et al., 2022). Possibly, the CPHQ may be better to measure dimensions of health beyond the initial PH scale, taking into account the context of persons.

### Methodological considerations

A strength of this study is the combination of both a data-driven and citizen-driven approach in a representative Dutch sample and the embeddedness in a theoretical framework of the Capability Approach. Previous tests of validity were empirically oriented (Doornenbal et al., 2022; Prinsen & Terwee, 2019). However, as also highlighted by Borsboom and colleagues, “validity cannot be solved by psychometric techniques or models alone.” (2004, p. 1062), it is needed to integrate multiple approaches from psychometrics, philosophy, and psychological theory. Therefore, we combined psychological theory, item construction with participants and experts, as well as comprehensive data analysis.

This study has some limitations. Different from the initial PH questionnaire, not all items of the CPHQ were formulated positively because we identified in the focus groups that vulnerable groups did not recognize themselves in items that were worded too positively. Inclusion of both positive and negative worded items has been suggested to reduce acquiescent response bias (Hinz et al., 2007) but others have shown the opposite (Sauro & Lewis, 2011). Although we did not find any indication that the reformulation affected the factorial structure and internal validity relative to the initial PH scale, further research is needed to test whether this also applies across other settings.

Further, we conducted factor analysis with many newly added items including those on context. Some of the initial items from the PH model (Van Vliet et al., 2021) did not remain in the final CPHQ questionnaire. For example, items of the “spiritual/existential domain” and “mental well-being” were not included in the CPHQ. This is most likely explained by the strong relationship between other factors such as enjoyment/contentment and the spiritual/existential domain (Doornenbal et al., 2022) and anxiety/depression (EQ-5D) and the CPHQ domains of relaxation, exclusion, social support, political representation, and health literacy. Thus, some items of the initial PH model appear to be replaced by related items.

At last, whereas the CPHQ explained more variance in resilience than the initial PH scale, the CPHQ still explained low variation in *autonomy* (ICECAP-A) and *self-care* (EQ-5D), which are factors important to the initial definition of Huber (i.e. “Health as the ability to self-manage”). This may be explained by the narrow scope of the measurements of both constructs. That is, *autonomy* was measured as “being able to be independent” (ICECAP-A) and *self-care* (EQ-5D) was measured as “being able to wash or clothe”. Both autonomy and self-care entail more than these measurements focus on. A*utonomy* in the context of health is defined as the right of people to make informed decisions about their medical care (Childers & Arnold, 2021). *Self-care* is often defined as the tasks performed at home by healthy people to prevent illness (Grady et al., 2014). These aspects were not included in the validation scales.

### Implications and future directions

In light of the previous work of Prinsen and Terwee (2019) and Huber et al. (2016), we showed that the initial PH measurement scale developed by Van Vliet et al. (2021) can be further advanced by incorporating personal, social, and environmental items derived from stakeholders and using the Capability Approach as a theoretical framework. The PH dialogue tool is very broad and includes aspects that either reflect health or influence health. This binary focus does not align well with the current paradigm in healthcare and health policy, which often focuses on traditional (disease) endpoints (i.e., “outcomes” in the field of epidemiology) and quality of life measurements.

The Capability Approach, which we chose as a theoretical framework, focuses less on traditional endpoints but more on people’s opportunities and capabilities towards such endpoints. Hence, by applying the Capability Approach, our measurement can help to develop and evaluate policies and other interventions according to their impact on people’s capabilities and not only on their actual functionings and feelings. Our measurement focuses on the extent to which people are able to feel healthy, and to what degree they have resources (e.g., the availability of healthy foods) needed for this capability, and to what extent conversions factors (e.g., living in a food desert) help transforming these resources into opportunities to feel well. Thus, our approach differs from the concept of PH, defined as “the ability to adapt and self-manage in the face of social, physical, and emotional challenges” (Huber et al., 2016). For that reason, we propose a refined definition of (Positive) health:

*”The extent to which one is capable to adapt and to thrive given one’s physical, mental, social and contextual opportunities”*

Further efforts are needed to test the reproducibility of the CPHQ, the responsiveness to change (i.e., to interventions), and the predictive validity (i.e., to biomedical indicators and healthcare utilization) to further test the construct validity and to create support for the use of CPHQ as measurement scale in healthcare and policy making.

## Conclusion

This study aimed to further develop and test a context-sensitive measurement of PH (CPHQ). By using a multimethodological approach, we advanced the initial PH questionnaire and added contextual items following the constitutive elements of the Capability Approach. The developed CPHQ showed adequate factorial validity and concurrent validity. Moreover, it accounts explicitly for resilience, which is one of the central elements of the concept of PH. Further research is needed to establish the relevance of self-management in de CPHQ and the reproducibility, responsiveness, and predictive validity of the measurement scale.

## Data Availability

All data produced in the present study are available upon reasonable request to the authors.

## Acknowledgment

We thank the citizens and experts for their contribution to the focus groups as well as the sounding board of the GEZOND METEN Consortium. We also thank Lise Beumeler and Mark Derks for their assistance in the data collection.

Funding for this project was received by the Fred Foundation, Noaber Foundation, and ZonMW.

## Supplementary material

**Supplemental Table 1.**
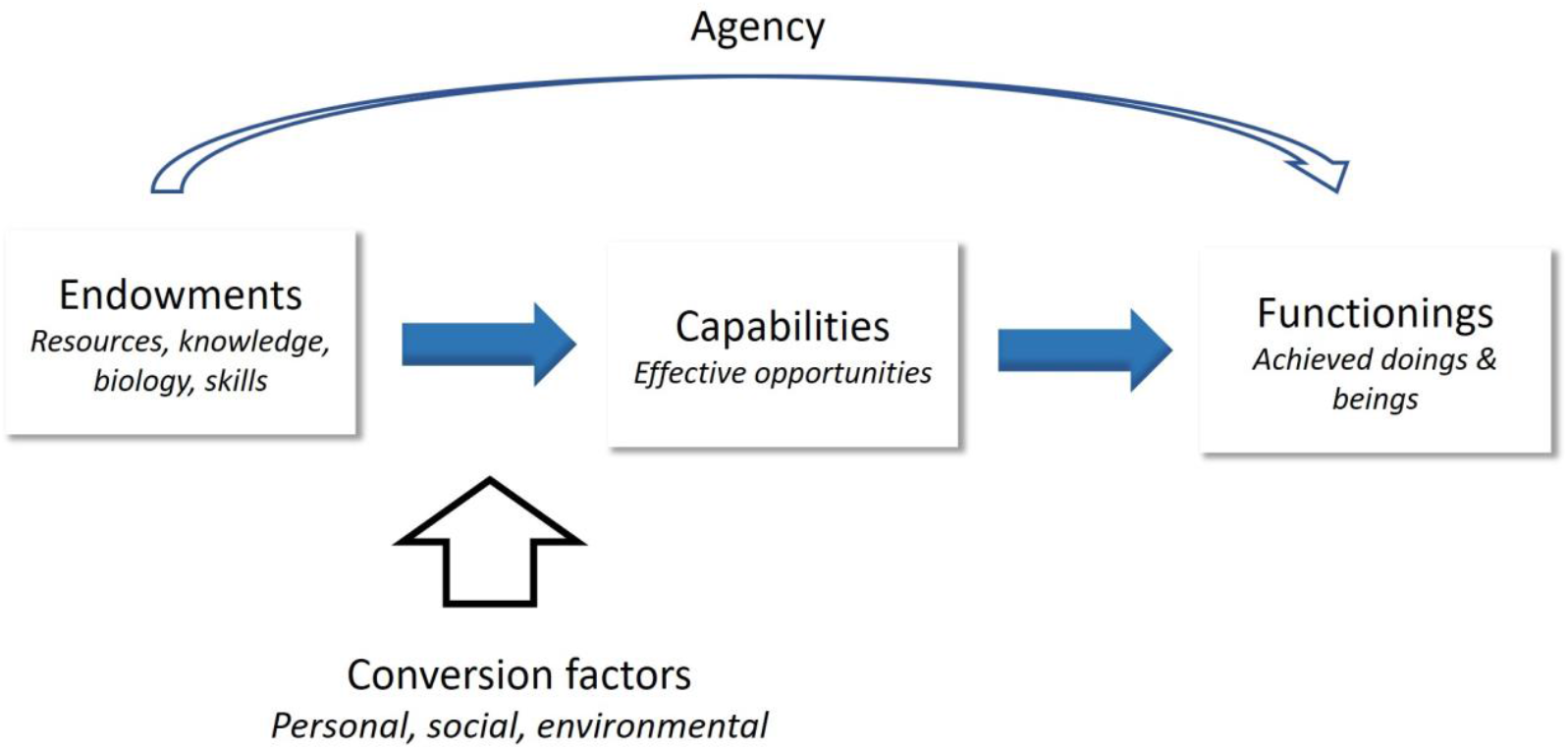
Central capabilities & Capability approach model, adapted from Chiappero-Martinetti & Venkatapuram, 2014. 1. Life 2. Bodily Health. 3. Bodily Integrity. 4. Senses, Imagination, and Thought 5. Emotions 6. Practical Reason. 7. Affiliation (living with or towards others, having social base). 8. Other Species. 9. Play. 10. Control over one’s Environment from a political and material perspective

**Supplemental Table 2.**
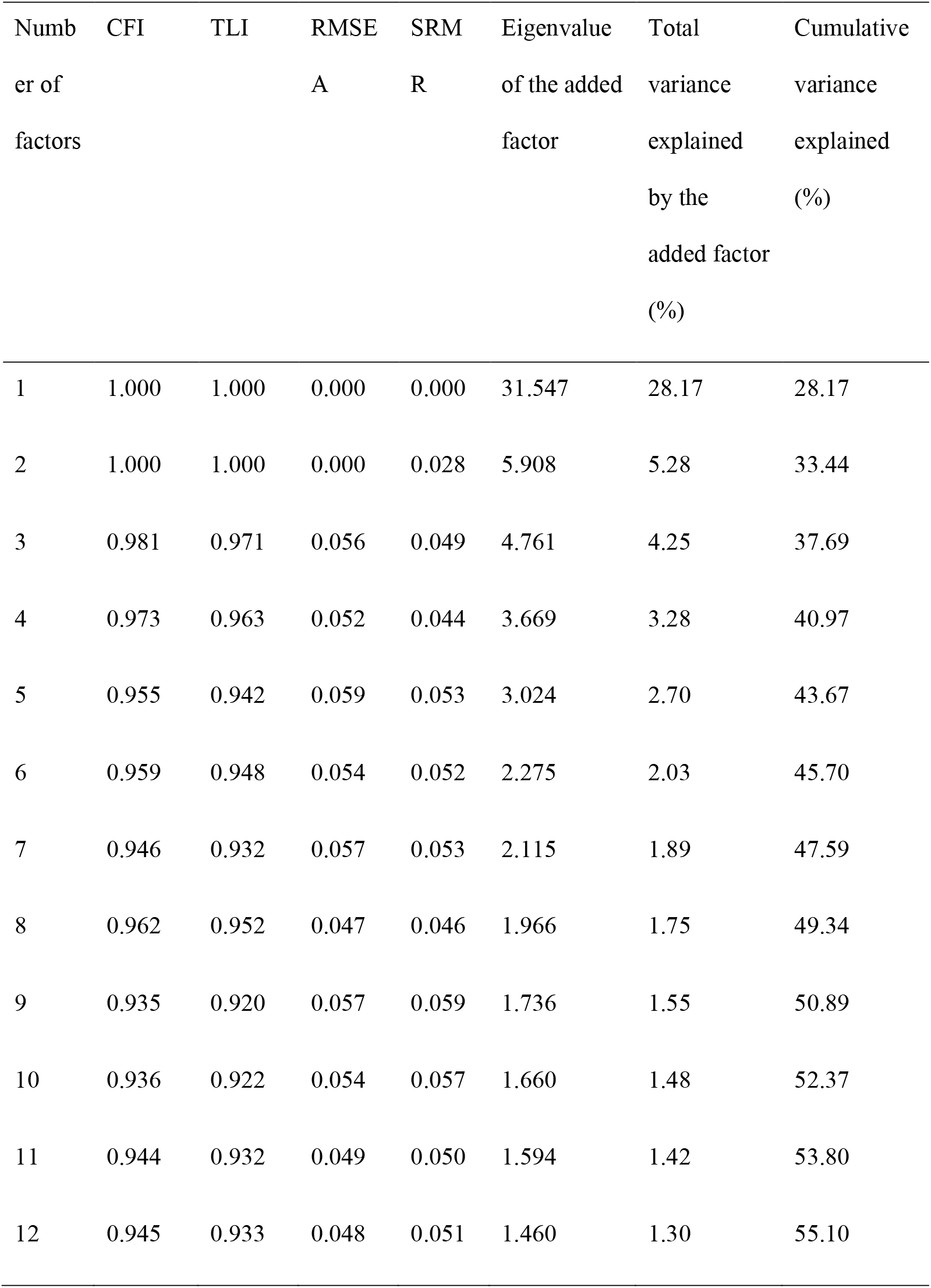
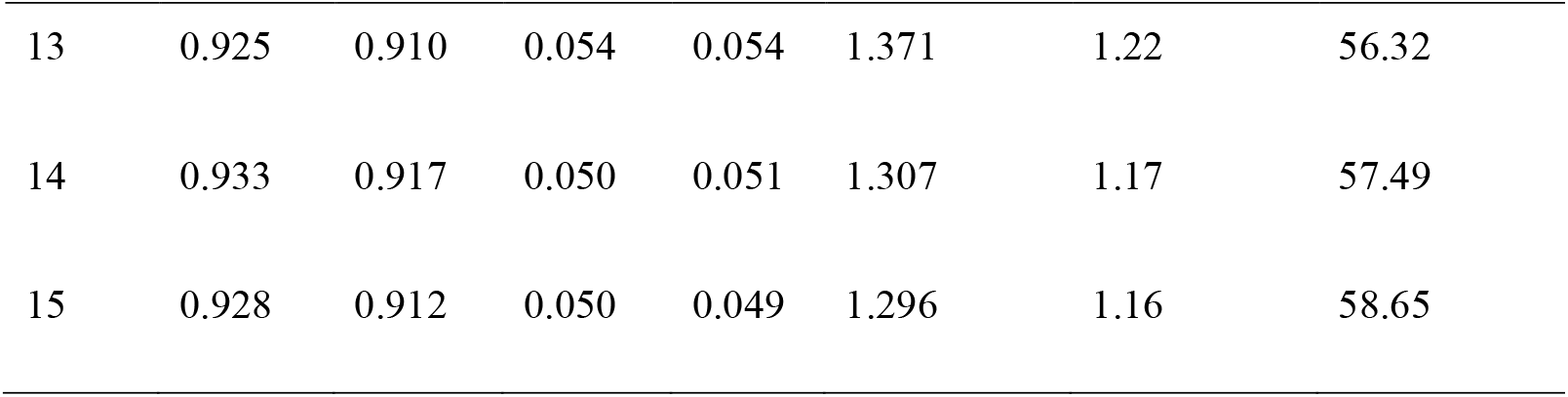
Model fit and variance explained for a series of exploratory factor analyses.

| Numb<br>er of<br>factors | CFI | TLI | RMSE<br>A | SRM<br>R | Eigenvalue<br>of the added<br>factor | Total<br>variance<br>explained<br>by the<br>added factor<br>(%) | Cumulative<br>variance<br>explained<br>(%) |
| --- | --- | --- | --- | --- | --- | --- | --- |
| 1 | 1.000 | 1.000 | 0.000 | 0.000 | 31.547 | 28.17 | 28.17 |
| 2 | 1.000 | 1.000 | 0.000 | 0.028 | 5.908 | 5.28 | 33.44 |
| 3 | 0.981 | 0.971 | 0.056 | 0.049 | 4.761 | 4.25 | 37.69 |
| 4 | 0.973 | 0.963 | 0.052 | 0.044 | 3.669 | 3.28 | 40.97 |
| 5 | 0.955 | 0.942 | 0.059 | 0.053 | 3.024 | 2.70 | 43.67 |
| 6 | 0.959 | 0.948 | 0.054 | 0.052 | 2.275 | 2.03 | 45.70 |
| 7 | 0.946 | 0.932 | 0.057 | 0.053 | 2.115 | 1.89 | 47.59 |
| 8 | 0.962 | 0.952 | 0.047 | 0.046 | 1.966 | 1.75 | 49.34 |
| 9 | 0.935 | 0.920 | 0.057 | 0.059 | 1.736 | 1.55 | 50.89 |
| 10 | 0.936 | 0.922 | 0.054 | 0.057 | 1.660 | 1.48 | 52.37 |
| 11 | 0.944 | 0.932 | 0.049 | 0.050 | 1.594 | 1.42 | 53.80 |
| 12 | 0.945 | 0.933 | 0.048 | 0.051 | 1.460 | 1.30 | 55.10 |
| 13 | 0.925 | 0.910 | 0.054 | 0.054 | 1.371 | 1.22 | 56.32 |
| 14 | 0.933 | 0.917 | 0.050 | 0.051 | 1.307 | 1.17 | 57.49 |
| 15 | 0.928 | 0.912 | 0.050 | 0.049 | 1.296 | 1.16 | 58.65 |

## Notes

### Competing Interest Statement

The authors have declared no competing interest.

### Author Declarations

Ethical approval was obtained from the Medical Ethical Review Board of the Leiden University Medical Center (protocol 19-035) and the Research Ethics Committee of the Faculty of Spatial Sciences, University of Groningen (protocol 202007).

